# Professional Identity and Its Influencing Factors Among Nursing Interns: The Mediating Role of Career Adaptability Between Social Support and Professional Identity

**DOI:** 10.64898/2026.10.14.26365571

**Authors:** Yanyan Zhao, Fuzhi Liu, Lanlan Chen, Xiuli Li, Biyu Wu

## Abstract

**Purpose:** The internship period is a critical stage for nursing students to develop professional values. This study aimed to investigate the current status of professional identity among nursing interns, analyze the effects of social support, perceived stress, psychological resilience, and career adaptability on professional identity, and explore the mediating role of career adaptability between social support and professional identity, so as to provide empirical evidence for enhancing professional identity and optimizing nursing talent training systems.

**Methods:** Using convenience sampling, 407 nursing interns from a tertiary hospital in Quanzhou were selected as participants from May to July 2025. Data were collected using a general information questionnaire, the Social Support Scale, the Perceived Stress Scale, the Psychological Resilience Scale, the Professional Identity Questionnaire for Nursing Students, the Career Adaptability Scale, and the Questionnaire for Nurse Intention to Retention Employed. SPSS 26.0 was used for reliability analysis, descriptive analysis, independent samples t-test, one-way ANOVA, Pearson correlation analysis, and multiple linear regression analysis. The PROCESS macro was employed to test the mediating effect.

**Results:** The total score of professional identity among nursing interns was (61.74±11.08), with 73.5% showing moderate professional identity. Parents’ attitude toward nursing, reasons for choosing the profession, personal attitude toward the major, and employment intention showed statistically significant differences in professional identity scores (p<0.05). Pearson correlation analysis revealed that professional identity was significantly positively correlated with social support (r=0.474), psychological resilience (r=0.389), and career adaptability (r=0.609) (p<0.01), and significantly negatively correlated with perceived stress (r=-0.378) (p<0.01). Multiple linear regression analysis indicated that career adaptability (β=0.309, p<0.001) and social support (β=0.383, p<0.001) were significant positive predictors of professional identity, whereas reasons for choosing the profession (β=-3.603, p=0.001) and employment intention (β=-7.116, p<0.001) were significant negative predictors. The model explained 49.2% of the variance in professional identity. Mediation effect analysis showed that career adaptability played a partial mediating role between social support and professional identity, with a mediation effect value of 0.288 (95% CI: 0.221–0.363), accounting for 40.2% of the total effect.

**Conclusions:** Nursing interns’ professional identity is at a moderately high level. Social support and career adaptability are positive influencing factors. Career adaptability plays a partial mediating role between social support and professional identity; social support indirectly enhances professional identity by strengthening career adaptability.

## 1. Introduction

Nursing shortages have become a global concern. By 2030, the worldwide deficit of nurses is projected to reach 4.5 million^[1]^’. A study based on 12.21 million hospitalized patients demonstrated that inadequate nurse staffing increases per capita nursing workload, leading to prolonged hospital stays, elevated inpatient mortality, and substantial economic burdens^[2]^. Compounding this issue, high turnover rates are prevalent among nursing personnel^[3]^. A meta-analysis of 21 studies across 14 countries found global nurse turnover rates ranging from 8% to 36.6%, with a pooled estimate of 16% (95% confidence interval: 14%–17%)^[4]^. The severe shortage and high turnover of nursing staff represent core challenges facing health systems, and retaining nursing graduates is a critical strategy for addressing this dilemma. As an essential reserve force for the future nursing workforce, graduates’ career choices are directly linked to the sustainable development of health services.

Professional identity is a critical psychological foundation during nursing interns’ transition from “student” to “nurse”, directly influencing their learning engagement and subsequent intention to stay, and ultimately helping mitigate high turnover and low employment rates^[5-6]^. During the internship phase, the encounter between clinical reality and professional ideals often causes notable fluctuations in professional identity, making it imperative to monitor the professional identity of this student cohort at this stage.

Professional identity is shaped by multiple interacting factors, including individual characteristics, behavioral habits, internship environments, psychological states, and occupational factors^[7–12]^. Among these, social support is widely recognized as a key external protective factor that promotes nursing students’ professional identity through positive feedback^[13]^. Career adaptability, as an important psychosocial resource for coping with occupational tasks and transitions, may play a significant mediating role between social support and professional identity^[14]^.

Although previous studies have examined the relationships between social support, career adaptability, and professional identity separately, empirical research integrating these three variables within a single mediation model and focusing specifically on nursing interns remains limited.Therefore, this study focuses on nursing interns, investigates their current status of professional identity, and constructs a mediation model along the core pathway of “social support

– career adaptability – professional identity”. Our findings aim to provide empirical evidence for enhancing professional identity among nursing interns and stabilizing the nursing workforce.

### Theoretical Framework

Social support was first systematically conceptualized by Cobb, who defined it as the emotional and material assistance individuals obtain from their social networks, such as family, friends, and mentors^[15]^. Building on this foundation, Cohen and Wills further distinguished two functional models: the main-effect model, which emphasizes the universal benefits of social support for physical and mental health, and the buffering-effect model, which posits that social support mitigates the adverse impact of stressful events^[16]^. In the career development domain, social support is regarded as a critical contextual resource that provides informational, emotional, and material security for individuals’ career exploration and decision-making^[17-18]^.

Career adaptability, derived from Savickas’s extension of Super’s career development theory, is defined as the psychosocial resources individuals possess to cope with current and anticipated occupational tasks, transitions, and traumas^[19]^. This construct encompasses four dimensions—concern, control, curiosity, and confidence—reflecting individuals’ capacity to actively shape their own careers^[20]^.

Professional identity refers to a clear and stable awareness of one’s vocational interests, abilities, and goals, representing a core psychological structure in career development. According to Holland’s theory, the formation of professional identity depends on the continuous integration of environmental feedback during career exploration, enabling individuals to adjust and confirm their self-positioning^[21]^. Existing research has primarily interpreted this formation mechanism through three theoretical lenses. First, social identity theory emphasizes that individuals construct self-concepts through membership in professional groups; positive emotional and value-based attachment to one’s occupational group strengthens professional identity^[22]^.Second, self-determination theory focuses on intrinsic motivation, positing that autonomy, competence, and relatedness are key psychological needs driving the formation of professional identity^[23]^. Third, social cognitive career theory (SCCT) integrates personal factors (e.g., self-efficacy), environmental factors (e.g., clinical atmosphere), and learning experiences, highlighting the dynamic interaction among individuals, environments, and behaviors. This framework offers a systematic analytical lens for understanding professional identity formation among specific populations such as nursing interns^[24]^.

### Hypothesis Development

A substantial body of empirical research has confirmed the significant positive effect of social support on professional identity. In a survey of 730 young professionals across 12 tertiary hospitals in China, perceived social support significantly influenced professional identity, with a total effect of b = 0.625^[25]^. Similarly, a study of 1,870 nursing interns confirmed that perceived social support positively affects professional identity^[26]^. Theoretically, this promoting effect can be explained by social cognitive career theory^[27-28]^, which posits that social support enhances individuals’ learning experiences, thereby fostering the development of vocational interests and career goals^[29–31]^. When individuals receive positive feedback and encouragement from significant others, their sense of value identification and belonging toward their chosen profession is strengthened accordingly^[32–34]^.

The development of career adaptability depends on resources individuals acquire from their environment^[35]^. A qualitative study among newly graduated nurses found that enhancing their adaptability and professional accomplishment, alongside improving the work environment, can effectively elevate societal recognition of the nursing profession and thereby stabilize the nursing workforce^[36]^. Furthermore, a study based on a self-management social cognitive career theory model examined the relationship between multi-source support and adolescent adaptability, demonstrating that family, peer, and teacher support directly influence adolescents’ career adaptability^[37]^. Thus, social support exerts a positive effect on career adaptability.

Career adaptability promotes professional identity through three primary pathways. First, career adaptability acts as a psychological transformer that converts cognitive resources into professional identity. Research among Turkish teachers found that cognitive control and flexibility had limited direct effects on professional identity; instead, their influence was fully mediated through the sequential pathway of career adaptability, career optimism, and career engagement, indicating that cognitive flexibility must operate via career adaptability to shape professional identity^[38]^. Second, career adaptability alleviates occupational stress and anxiety, creating psychological space for the cultivation of professional identity. A longitudinal study of nursing students found that pre-internship perceived stress significantly predicted post-internship anxiety (β = 0.474, p < 0.001), whereas career adaptability significantly attenuated this effect (β = −0.009, p < 0.01), with the buffering effect being stronger among those with higher adaptability^[39]^. Third, career adaptability and professional identity form a mutually reinforcing dynamic cycle. Studies of Chinese engineering students and pre-service kindergarten teachers confirmed that professional identity positively influences career adaptability, with learning engagement, teaching efficacy, and self-leadership serving as mediators or chain mediators^[40-41]^. Individuals with high professional identity tend to invest more in professional practice, thereby enhancing career adaptability; in turn, high adaptability strengthens individuals’ capacity to meet challenges and sustain professional engagement, further consolidating professional identity. Conversely, research among clinical nurses demonstrated that organizational dehumanization was negatively associated with career adaptability (total effect = −0.357, p < 0.001), with self-compassion partially mediating this relationship and psychological safety buffering the negative indirect effect at high levels, suggesting that supportive environments can indirectly preserve professional identity by protecting adaptability^[42]^.

Collectively, these findings establish the mediating mechanism of career adaptability between social support and professional identity. The underlying logic is that social support, as an external resource, enhances individuals’ internal psychosocial resources for coping with occupational tasks and transitions (i.e., career adaptability), which in turn promotes their identification with and internalization of their profession^[43-44]^. This perspective aligns with conservation of resources (COR) theory, which posits that external support helps individuals accumulate key psychological resources, thereby fostering positive occupational psychological states^[45]^.

Based on the above theoretical and empirical evidence, this study proposes the following hypotheses (see Figure 1):

**H1:** Social support has direct positive effects on both career adaptability and professional identity.

**H2:** Social support indirectly affects professional identity through the mediating role of career adaptability.

**Figure. 1.**
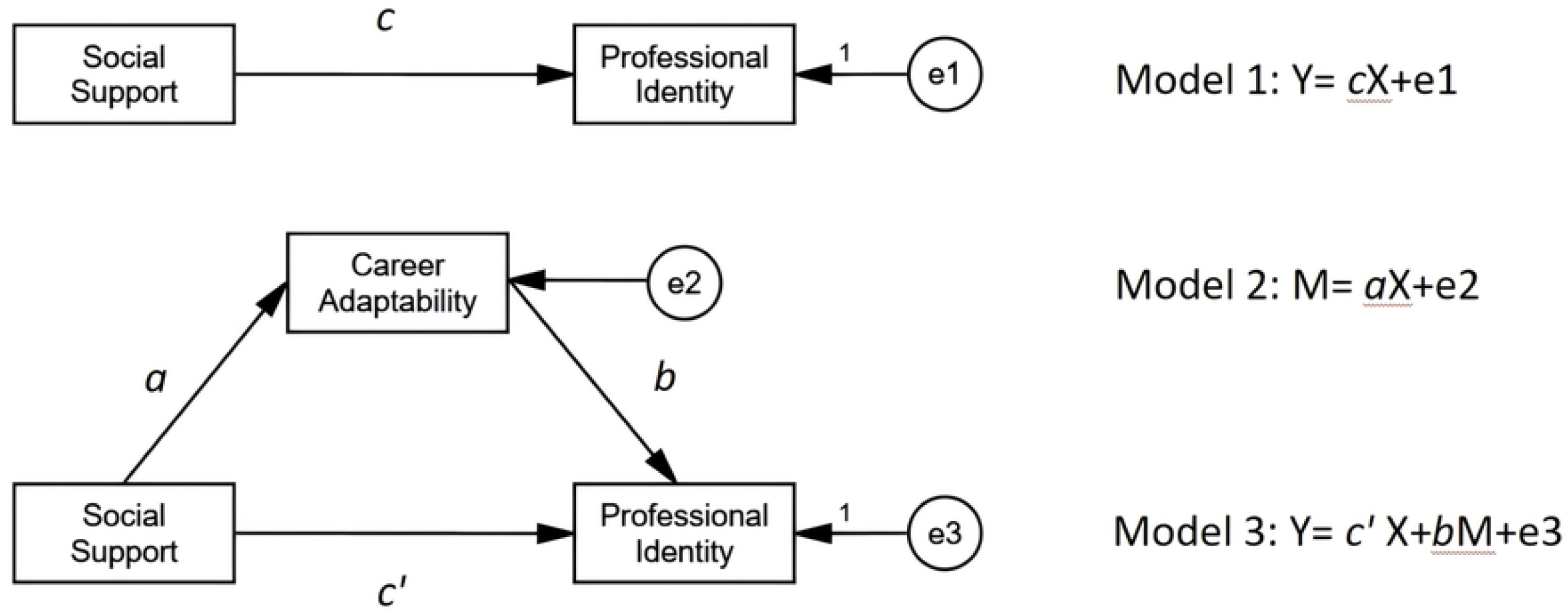

**Figure. 2.**
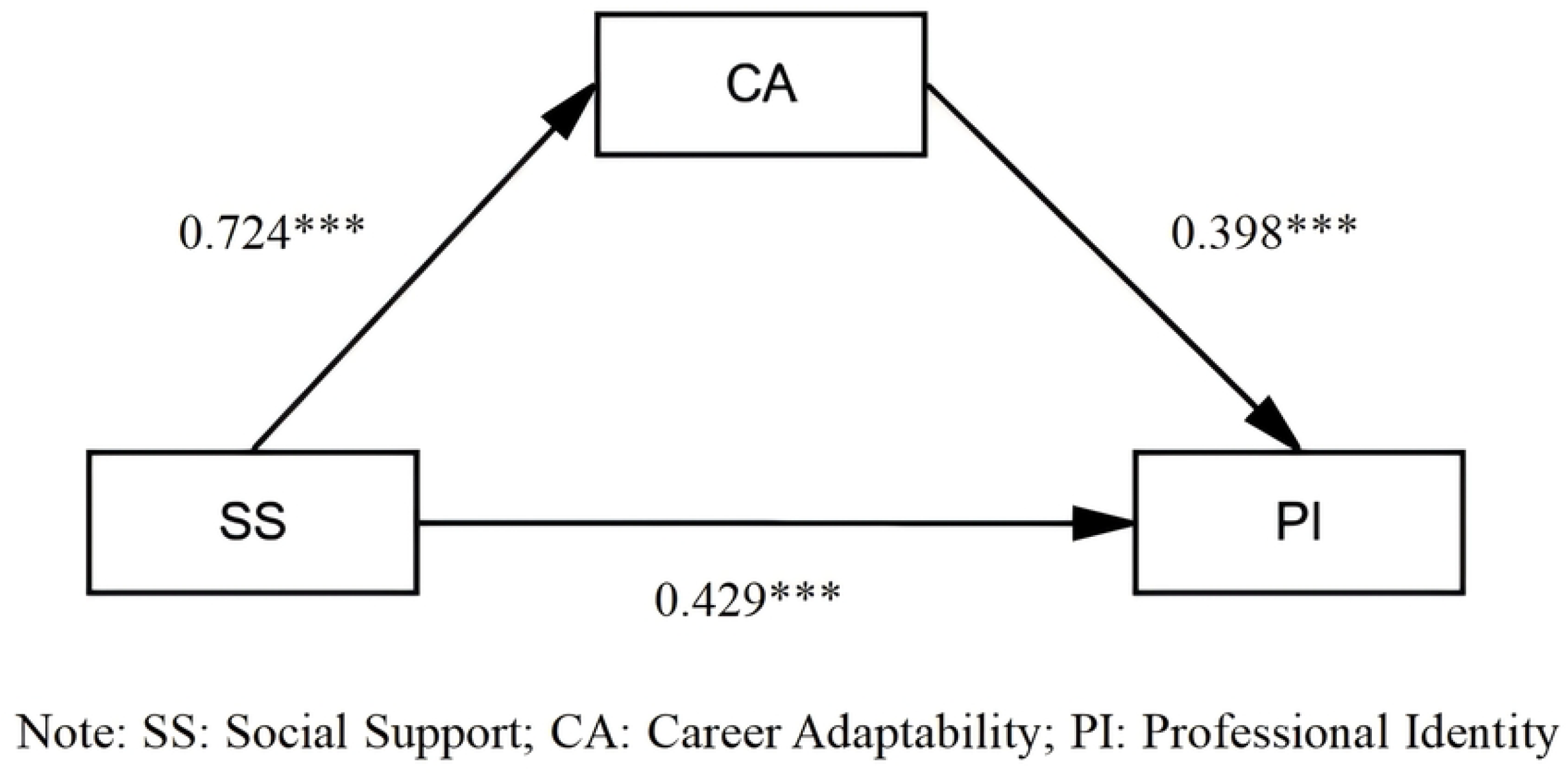

## 2. Materials and methods

### 2.1 Participants

The study recruited nursing interns at Quanzhou First Hospital. Inclusion criteria included being an officially enrolled nursing student currently undertaking clinical nursing internship, having full civil capacity to independently understand and complete the questionnaire, and providing voluntary participation with consent for data use limited to academic research. Students were excluded if they were from non-nursing majors or had not yet entered the clinical internship phase; if their internship had been suspended for over one month due to leave (personal or medical) without resumption; if they had cognitive impairment or psychiatric disorders precluding normal questionnaire completion; or if they declined participation or failed to provide informed consent.

### 2.2 Measures

#### Basic Demographic Information Questionnaire

A self-designed general information questionnaire was developed, covering three dimensions: (1) basic demographic characteristics, including age, gender, education, and family background; (2) lifestyle behaviors, including smoking, alcohol consumption, and physical exercise; and (3) factors related to professional identity, including parents’ attitude toward nursing, reasons for choosing the nursing major, and employment intention.

#### Social Support

The Social Support Rating Scale was developed by Xiao Shuiyuan^[46]^. It consists of 10 items across three dimensions: objective support, subjective support, and utilization of support. Items 1– 4 and 8–10 are rated on a 4-point scale, Item 5 is rated on a 5-point scale, and Items 6–7 are multiple-choice items analyzed qualitatively only. The total score ranges from 12 to 66, with higher scores indicating greater social support. The classification criteria are as follows: 12–33 indicates low support, 34–45 indicates moderate support, and 46–66 indicates high support. In the present study, the Cronbach’s α coefficient of this scale was 0.754.

#### Stress

The Perceived Stress Scale was originally developed by Cohen et al. in 1983^[47]^. The Chinese version was revised by Yang Tingzhong et al. in 2003^[48]^. It is used to assess an individual’s perceived stress level over the past month. The scale comprises 14 items across two dimensions: tension and lack of control. Items 4, 5, 6, 7, 9, 10, and 13 are reverse-scored. A 5-point Likert scale is used (0 = never, 4 = always). The total score ranges from 0 to 56, with higher scores indicating greater perceived stress. In the present study, the Cronbach’s α coefficient of this scale was 0.894.

#### Resilience

The Connor-Davidson Resilience Scale is an abbreviated version extracted by Campbell-Sills and Stein in 2007 from the original 25-item scale^[49]^. The Chinese version was revised by Yu Xiaonan, Zhang Jianxin et al. in 2007^[50]^. It is used to assess an individual’s adaptability and resilience in adversity. The scale consists of 10 items with a unidimensional structure, rated on a 5-point Likert scale (0 = not at all true, 4 = true nearly all the time). The total score ranges from 0 to 40, with higher scores indicating greater resilience. In the present study, the Cronbach’s α coefficient of this scale was 0.972.

#### Retention Intention

The Questionnaire for Nurses’ Intention to Remain Employed is used to assess nurses’ intention to remain in their positions^[51-52]^. The questionnaire comprises 6 items covering six aspects: professional identity, likelihood of remaining in the position, turnover intention, job satisfaction, career development, and emotional attachment. Item 3 is reverse-scored. A 5-point Likert scale is used (1 = completely disagree, 5 = completely agree). The total score ranges from 6 to 30, with higher scores indicating stronger intention to remain. In the present study, the Cronbach’s α coefficient of this questionnaire was 0.787.

#### Career Adaptability

The Career Adaptability Scale is used to assess an individual’s adaptability in the occupational environment^[53]^. This scale, as translated by Hou, is a 24-item instrument organized into four dimensions (six items each). It employs a 5-point Likert scoring system, generating composite scores from 24 to 120, wherein elevated scores reflect enhanced career adaptation ability^[54]^. In the present study, the Cronbach’s α coefficient of this scale was 0.977.

#### Professional Identity

The Professional Identity Scale was developed by Hao Yufang^[55]^. It is the most widely used scale with established reliability and validity specifically designed for nursing students in China. The scale comprises 17 items across five dimensions: professional self-concept (6 items), benefits of staying and risks of leaving (4 items), social support and self-reflection (3 items), autonomy in career choice (2 items), and social persuasion (2 items). Item 15 is reverse-scored. A 5-point Likert scale is used (1 = strongly disagree, 5 = strongly agree). The total score ranges from 17 to 85, with higher scores indicating stronger professional identity. The classification criteria are as follows: 17–42 indicates low identity, 43–68 indicates moderate identity, and 69–85 indicates high identity. In the present study, the Cronbach’s α coefficient of this scale was 0.950.

### 2.3 Procedures

During the pre-internship orientation period from May to July 2025, we provided centralized explanations to all participants regarding the purpose of this study and the procedures for completing the questionnaire. This study was conducted in strict accordance with the Declaration of Helsinki and received formal approval from the Institutional Review Board (Approval No. Quan Yi Lun 2025K163).

A self-administered online electronic questionnaire was created using the Wenjuanxing platform. An informed consent form was attached at the beginning of the questionnaire, and participants were required to read and provide consent before proceeding. Prior to distribution, we verbally restated the research purpose and feedback procedures, and explicitly informed participants that all collected data would be used solely for scientific research and that no personal information would be disclosed.

The questionnaire link was disseminated through social media platforms such as WeChat and QQ, specifically via nursing internship groups and university class groups. Each participant was restricted to a single submission. The estimated minimum completion time was 5 minutes, and responses falling below this threshold were excluded. The research team monitored daily response rates and promptly addressed any questions raised by participants during completion to ensure a smooth data collection process.

A total of 415 questionnaires were distributed, all 415 were returned, yielding a response rate of 100.0%. After data verification and cleaning, 8 invalid questionnaires (including those with insufficient response times) were excluded. Ultimately, 407 valid questionnaires were obtained, resulting in a valid response rate of 98.1%.

### 2.4 Data Analysis

Data were analyzed using SPSS version 26.0 and the PROCESS macro developed by Hayes. Descriptive statistics were computed, with categorical variables expressed as frequencies and percentages (%), and continuous variables as mean ± standard deviation. Between-group differences were assessed using independent-samples t-tests for two-group comparisons or one-way analysis of variance (ANOVA) for multi-group comparisons, while Pearson correlation analysis was performed to examine bivariate associations among variables. To identify factors influencing professional identity, multiple linear regression with the enter method was conducted with professional identity as the dependent variable, and tolerance values and variance inflation factors (VIF) were examined for all independent variables to evaluate multicollinearity. Mediation analysis was performed using the PROCESS macro (Model 4) to test the mediating role of career adaptability in the relationship between social support and professional identity, with the bootstrap method applied using 5,000 resamples to generate bias-corrected 95% confidence intervals (CIs) for the indirect effects. All statistical tests were two-tailed, and a P-value < 0.05 was considered statistically significant.

## 3.Results

### 3.1 Demographic Characteristics of the Participants

Among the 407 nursing interns, females accounted for 88.5% (n=360), with a mean age of 20.56±0.69 years. The majority had a college degree or below (89.7%, n=365), while 10.3% (n=42) held an undergraduate degree. Most participants came from village areas (64.4%), were not the one child in the family (85.7%), and were from non-single-parent families (92.1%). Parental support for the nursing profession was reported by 97.8% (n=398) of the respondents. Regarding the reasons for choosing nursing, teacher or parent recommendation was the primary factor (62.2%), followed by personal interest (32.1%). In terms of personal attitude toward the nursing major, 47.1% expressed “favorable,” 50.9% indicated “moderate” attitude, and 93.6% were willing to engage in nursing work in the future. With respect to lifestyle habits, smokers and alcohol drinkers accounted for 1.2% and 0.5%, respectively; the majority of participants reported “sometimes” physical exercise (67.4%) (see Table 1).

### 3.2Analysis of Influencing Factors of Professional Identity

#### 3.2.1 Distribution of Professional Identity among Nursing Interns with Different Characteristics

The results showed that professional identity scores differed significantly across groups defined by education, parental attitude, reason for major selection, attitude toward major, employment intention, and physical activity (all *P* < 0.05). No significant differences were observed for the remaining variables. Detailed results are presented in Table 2.

#### 3.2.2 Correlation Analysis between Major Variables and Professional Identity

Pearson correlation analysis was conducted among professional identity, social support, stress, resilience, career adaptability, and retention intention. The results indicated that professional identity was significantly and positively correlated with social support (*r* = 0.474), resilience (*r* = 0.389), and career adaptability (*r* = 0.609) (all *P* < 0.01), whereas it was significantly and negatively correlated with stress (*r* = −0.378) (*P* < 0.01). Using the cutoff points of 17–42, 43–68, and 69–85 for low, moderate, and high professional identity, respectively, the mean score of the participants was 61.74 ± 11.08, indicating a moderately high level. These results are presented in Table 3.

### Multiple Linear Regression Analysis of Factors Influencing Professional Identity among Nursing Interns

Multiple linear regression using the enter method was performed with professional identity as the dependent variable. Independent variables included those with P < 0.20 in univariate analysis and variables reported as statistically significant in previous literature (age, gender, education, parental attitude, reason for major, employment intention, attitude toward the major, and physical exercise), psychological scales (social support, perceived stress, and resilience), and occupational scales (career adaptability and retention intention). Age and sex were adjusted for in the model. The results are presented in Table 4.

The regression model was statistically significant overall (*F* = 66.643, *P* < 0.001), with a coefficient of determination (*R*^2^) of 0.500 and an adjusted *R*^2^ of 0.492, indicating that the model explained 49.2% of the variance in professional identity among nursing interns. The variance inflation factor (VIF) values for all independent variables were less than 2, suggesting no multicollinearity concerns and good model fit.

The results showed that career adaptability (*β* = 0.309, *P* < 0.001) and social support (*β* = 0.383, *P* < 0.001) were significantly and positively associated with professional identity, whereas reason for major selection (*β* = −3.603, *P* < 0.001) and employment intention (*β* = −7.116, *P* < 0.001) were significantly and negatively associated with professional identity.

### 3.4 Mediation Analysis of Career Adaptability between Social Support and Professional Identity

To examine whether career adaptability mediated the association between social support and professional identity, Model 4 of the Hayes’ PROCESS macro for SPSS was employed. Social support was entered as the independent variable (X), professional identity as the dependent variable (Y), and career adaptability as the mediator (M). The bootstrap procedure was performed with 5,000 resamples to generate bias-corrected 95% confidence intervals (CIs).

#### 1. Path coefficient analysis

All path coefficients in the mediation model were statistically significant (all P < 0.001), indicating that all proposed pathways were supported. See Table 5 for details.

#### 2. Bootstrap test of the mediation effect

The bootstrap results (5,000 resamples) revealed a significant indirect effect of social support on professional identity via career adaptability, with the 95% CI excluding zero. The indirect effect was 0.288, accounting for 40.2% of the total effect. Detailed results are presented in Table 6.

## 4.Discussion

### 4.1Current Status of Professional Identity Among Nursing Interns

The present study found that the professional identity of nursing interns was at a moderate-to-high level (61.74 ± 11.08), which is largely consistent with the findings of similar domestic studies^[56]^. Notably, 73.5% of the participants scored at a moderate level of professional identity, indicating that, for the majority of nursing interns undergoing the transition from student to nurse, professional identity has not yet been fully consolidated and remains highly malleable.

This result is broadly in line with previous research. A prospective cohort study reported that a relatively high proportion of nursing interns exhibited a moderate level of professional identity, a distributional pattern consistent with that observed in our study^[57]^. However, a cross-sectional survey^[58]^ reported a slightly lower professional identity score among nursing students (56.18 ± 8.75) than that found in the present study, a discrepancy that may be attributable to differences in sample sources (e.g., geographic regions and hospital grades) and in the measurement instruments employed. It is also noteworthy that only 3.2% of the participants in our study demonstrated a low level of professional identity. This may be related to the high proportion of participants whose parents supported their choice of nursing as a major (97.8%), as family support is an important protective factor for professional identity^[59]^ and may, to some extent, buffer against its negative development.

Univariate analysis in this study showed that parental attitudes, reasons for choosing the nursing major, personal attitudes toward the profession, and willingness to pursue nursing employment had statistically significant effects on professional identity (p < 0.05). Participants whose parents supported their choice scored significantly higher on professional identity (62.10 ± 10.83) than those whose parents did not (45.89 ± 11.03); those who chose nursing out of personal interest scored significantly higher (67.98 ± 10.16) than those who were assigned to the major through transfer allocation (49.74 ± 11.08); and those who reported liking the nursing profession scored significantly higher (68.36 ± 9.36) than those who did not (35.00 ± 7.09). These findings corroborate the “autonomy support principle” in the formation of professional identity—when individuals choose an occupation based on intrinsic motivation rather than external pressure, their level of identification is higher—which is highly consistent with the core propositions of self-determination theory^[60–61]^. In addition, the present study found that physical exercise was closely associated with professional identity: the higher the frequency of exercise, the higher the professional identity score, a pattern consistent with previous reports^[62–63]^. It is worth noting that demographic variables such as gender, age, and place of origin showed no significant differences in the univariate analysis. This may be related to the high homogeneity of the sample (88.5% female, with ages concentrated at 20–21 years) and also suggests that, among nursing interns, psychosocial factors (social support: r = 0.474, p < 0.01; perceived stress: r = −0.378, p < 0.01; psychological resilience: r = 0.389, p < 0.01) may have greater explanatory power than demographic characteristics.

To further identify the key determinants of professional identity, a multiple linear regression model was constructed after controlling for demographic variables. The results showed that career adaptability (β = 0.309, p < 0.001) and social support (β = 0.383, p < 0.001) exerted significant positive effects on professional identity, whereas the reason for choosing the nursing major (β = −3.603, p = 0.001) and employment intention (β = −7.116, p < 0.001) exerted significant negative effects. Together, these variables explained 49.2% of the variance in professional identity.

### 4.2 Career Adaptability: An Internal Driving Force in the Construction of Professional Identity

In this study, career adaptability was the strongest factor influencing the professional identity of nursing interns. The magnitude of this effect (standardized β = 0.390) indicates that career adaptability does not merely reflect passive adaptive capacity; rather, it functions as an “engine” that actively drives the construction of professional identity. When nursing interns possess higher levels of career adaptability—encompassing the dimensions of concern, control, curiosity, and confidence—they are more inclined to engage in proactive learning behaviors, integrate actively into the clinical environment, and accumulate positive professional experiences, thereby strengthening their value recognition of and emotional commitment to the nursing profession^[64]^. This finding is consistent with previous research and extends our understanding of the pivotal role of career adaptability in nursing education. Wang et al. reported that a dual-mentorship model significantly enhanced both career adaptability and professional identity among nursing students—two interrelated competencies regarded as essential for successful career construction^[65]^. Similarly, prior research has shown that career adaptability significantly predicts professional identity over time among Chinese undergraduate nursing students, with more adaptable individuals tending to hold clearer career goals and stronger professional commitment^[66]^.

### 4.3 Social Support: A Critical External Resource for the Formation of Professional Identity

In this study, social support exerted a significant positive effect on the professional identity of nursing interns (standardized β = 0.253), confirming that support from family, peers, and the clinical team constitutes a critical external resource for the formation of professional identity. This finding is consistent with a substantial body of existing evidence demonstrating a positive association between nursing students’ professional identity and the level of social support they receive from various sources^[67]^.

The theoretical basis for this relationship can be traced to Social Cognitive Career Theory (SCCT), which posits that contextual supports—including social support provided by significant others—enhance individuals’ learning experiences and, in turn, foster the development of vocational interests and career goals^[68]^. When nursing students receive positive feedback, encouragement, and instrumental assistance from key figures in their professional and personal networks, their perceived value of and sense of belonging to the nursing profession are correspondingly strengthened^[69]^. The univariate analysis of the present study also confirmed this pattern: participants with parental support scored significantly higher on professional identity (62.10 ± 10.83) than those without (45.89 ± 11.03) (p < 0.001).

Notably, the reason for choosing the nursing major and employment intention showed significant negative effects in the regression model, revealing individual differences in professional identity. Compared with those who chose nursing out of “personal interest,” participants who entered the major on the “recommendation of teachers or parents” or through “institutional transfer allocation” obtained lower professional identity scores; likewise, participants who were unwilling to pursue nursing employment scored significantly lower than those who were willing. These results are consistent with the predictions of self-determination theory, suggesting that intrinsic motivation (e.g., personal interest) serves as an important driving force in the formation of professional identity, whereas passive, externally driven choices may weaken students’ identification with the profession—that is, when intrinsic motivation is lacking, the formation of professional identity may be constrained even in the presence of external support^[70–71]^. In the multiple linear regression analysis, a model including only social support and career adaptability already achieved a high level of explanatory power for professional identity (adjusted R^2^ = 0.439), with only a limited increase after additional variables were entered (adjusted R^2^ = 0.492). Furthermore, the moderate level of social support (36.50 ± 7.32), together with the high level of career adaptability (109.35 ± 13.97), may jointly contribute to a moderate level of professional identity (61.74 ± 11.08) among nursing interns. Taken together, these results indicate that social support and career adaptability are the principal explanatory variables of professional identity among nursing interns, with career adaptability making the more prominent contribution.

### 4.4 The Mediating Role of Career Adaptability: Integrating Internal and External Resources

To further elucidate the relationships among social support, career adaptability, and professional identity, this study constructed a mediation model of “social support → career adaptability → professional identity.” The mediation analysis showed that social support had a significant positive effect on career adaptability (β = 0.724, p < 0.001) and that career adaptability had a significant positive effect on professional identity (β = 0.398, p < 0.001), supporting Hypothesis 1. After the mediating variable was introduced, the direct effect of social support on professional identity remained significant (0.429, p < 0.001), indicating that career adaptability played a partial mediating role and thus supporting Hypothesis 2. The indirect effect was 0.288, accounting for 40.2% of the total effect, with a bootstrap 95% confidence interval of (0.221, 0.363). These results reveal that social support not only directly enhances nursing students’ professional identity but also exerts an indirect effect by improving their career adaptability, with the indirect pathway accounting for nearly half of the total effect.

At the theoretical level, this finding provides new empirical support for the Conservation of Resources (COR) theory and illuminates the underlying mechanism of career adaptability as an “empowering mediator.” According to COR theory, individuals are inherently motivated to acquire, retain, and protect valued resources; external support (such as social support from family, school, and the clinical environment) functions as a contextual resource that enables individuals to accumulate internal psychological resources (such as career adaptability), which in turn facilitate the formation of positive occupational psychological states (such as professional identity)^[72]^.

The present study further integrates Career Construction Theory with COR theory to form an explanatory chain of “resource infusion—capacity transformation—identity internalization": social support provides the “resources for adaptation,” career adaptability represents the “capacity for adaptation,” and together they ultimately yield the “outcome of adaptation"—professional identity. Recent empirical evidence supports this integrative perspective: in a study of 4,252 Chinese first-year university students, parental career support was found to enhance career identity both directly and indirectly through career adaptability and career construction behaviors (a sequential mediation of “adaptability → behavior → identity”), highlighting how external support activates core adaptive processes that foster the formation of career identity^[73]^. For nursing interns, this pathway can be conceptualized as a “resource caravan"—external support resources (e.g., encouragement from clinical preceptors, emotional support from family, and peer camaraderie) are converted into internal adaptive resources that, in turn, drive the process of identity construction^[74]^.

### 4.5 Integrated Discussion and Mechanism Synthesis

Taken together, the formation of professional identity among nursing interns follows a progressive pathway of “external resources → internal resources → occupational psychology.” Social support, as an external contextual resource, provides the foundational guarantee for the formation of professional identity; career adaptability, as an internal psychological resource, serves as the key mediator through which external support is transformed into professional identity; and choice motivation and employment intention modulate this process at the level of individual agency. Integrating the regression and mediation models, career adaptability emerges both as the strongest determinant of professional identity (standardized β = 0.390) and as the central transmission pathway through which social support exerts its influence (with the indirect effect accounting for 40.2% of the total effect). This dual role positions career adaptability as the hub connecting external support with internal identity, underscoring its central place in the professional psychological development of nursing interns. In addition, the association between intention to stay and professional identity was weak in this study (r = 0.029), which may be related to the particularities of the internship stage—before formally entering professional practice, interns’ judgments about remaining in the profession are based more on imagination than on authentic occupational experience. Future longitudinal studies are warranted to examine the extent to which professional identity formed during the internship period predicts retention behavior at the early career stage.

## 5. Limitations

Several limitations should be acknowledged in interpreting and generalizing the findings of this study, arising from the design and implementation of the methodology.

First, in terms of sampling, a convenience sampling method was employed, and participants were recruited solely from one tertiary hospital in Quanzhou. This single-site recruitment may introduce selection bias and limit the generalizability of the results.

Second, regarding study design, this was a cross-sectional survey in which all variables were measured at a single time point, precluding any inferences about causal or temporal relationships. Although the mediation model was constructed based on theoretical assumptions, the dynamic causal pathways among social support, career adaptability, and professional identity require further validation through longitudinal study designs.

Third, concerning common method bias, all data were derived from self-reported questionnaires, and the observed relationships among variables may be influenced by common method variance. Fourth, with respect to variable selection and measurement, although core variables such as social support, perceived stress, and career adaptability were included in this study, other potentially important variables—such as internship environmental factors (e.g., teaching quality, ward climate) and individual factors (e.g., self-efficacy, achievement motivation)—were not fully incorporated into the model, leaving room for improvement in explanatory power.

Fifth, regarding sample representativeness, the sample was predominantly female (88.5%), and the majority held associate degrees or below (89.7%). Although this distribution is generally consistent with the overall population of nursing interns in China, caution is warranted when generalizing the findings to populations with more balanced gender distributions or those with bachelor’s or higher degrees.

Future research could adopt multi-center stratified random sampling methods to expand the coverage and diversity of the sample; conduct longitudinal follow-up studies to delineate causal pathways among variables; incorporate objective indicators or multi-source data to reduce common method bias; and explore additional potential mediators and moderators to further refine the theoretical model of professional identity formation among nursing interns.

## 6. Conclusions

Nursing interns had a mean professional identity score of 61.74 ± 11.08, indicating a moderately high level; 73.5% of the participants reported moderate professional identity. Social support (*β* = 0.383) and career adaptability (*β* = 0.309) were positively associated with professional identity, whereas reason for major selection (*β* = −3.603) and employment intention (*β* = −7.116) were negatively associated. Mediation analysis revealed that career adaptability partially mediated the effect of social support on professional identity (indirect effect = 0.288, accounting for 40.2% of the total effect). Specifically, social support not only directly enhanced professional identity but also indirectly promoted it by strengthening career adaptability.

### Supporting information

S1 Fig. Conceptual framework of the hypothesized mediation model linking social support, career adaptability, and professional identity.

S2 Fig. Structural equation model with standardized path coefficients for the mediating effect of career adaptability.

S1. Raw Data

## Data Availability

All relevant data are within the manuscript and its Supporting Information files.

## Glossary

SSRS: Social Support Rate Scale
SCCT: Social Cognitive Career Theory
COR: Conservation of Resources Theory
CI: Confidence Interval
SD: Standard Deviation
VIF: Variance Inflation Factor
ANOVA: Analysis of Variance

## Acknowledgments

We thank the participantes in the study and acknowledge the reviewers and editors for viewing our manuscript.

